# Impact of MEK inhibitor therapy on neurocognition in neurofibromatosis type 1

**DOI:** 10.1101/2020.12.18.20248334

**Authors:** Karin S. Walsh, Pamela L. Wolters, Brigitte C. Widemann, Allison A. del Castillo, Maegan D. Sady, Tess Inker, Marie Claire Roderick, Staci Martin, Mary Anne Toledo-Tamula, Kari Struemph, Iris Paltin, Victoria Collier, Kathy Mullin, Michael J. Fisher, Roger J. Packer

**Affiliations:** Children’s National Hospital; George Washington University School of Medicine; National Cancer Institute/NIH; Children’s Hospital of Philadelphia; University of Pennsylvania Perelman School of Medicine; Clinical Research Directorate, Frederick National Laboratory for Cancer Research

## Abstract

**Objective:** NF1-associated cognitive impairments carry significant life-long morbidity. The lack of targeted biologic treatments remains a significant unmet need. We examine changes in cognition in patients with NF1 in the first 48 weeks of MEK inhibitor (MEKi) treatment.

**Methods:** 59 NF1 patients ages 5-27 on a MEKi clinical trial treating plexiform neurofibroma underwent pre-treatment and follow-up cognitive assessments over 48-weeks of treatment. Performance tasks (Cogstate) and observer-reported functioning (BRIEF) were primary outcomes. Group-level (paired *t*-tests) and individual-level analyses (reliable change index; RCI) were used.

**Results:** Analysis showed statistically significant improvements on BRIEF compared to baseline (24-week BRI: *t*_(58)_=3.03, *p*=.004, *d*=0.24; 48-week MCI: *t*_(39)_=2.70, *p*=.01, *d*=0.27). RCI indicated more patients had clinically significant improvement at 48-weeks than expected by chance (*Chi Square*=11.95, *p*=.001, OR=6.3). Group-level analyses indicated stable performance on Cogstate (*p*>.05). RCI statistics showed high proportions of improved working memory (24-weeks *Chi Square*=8.36, *p*=.004, OR=4.6 and 48-weeks *Chi Square*=9.34, *p*=.004, OR=5.3) but not visual learning/memory. Patients with baseline impairments on BRIEF were more likely to show significant improvement than non-impaired patients (24-weeks 46% v. 8%; *Chi Square*=9.54, *p*=.008, OR=9.22; 48-weeks 63% v. 16%; *Chi Square*=7.50, *p*=.02, OR=9.0).

**Interpretation:** Our data shows no evidence of neurotoxicity in 48-weeks of treatment with a MEKi and a potential clinical signal supporting future research of MEKi as a cognitive intervention.

## Introduction

Learning and cognitive deficits are highly prevalent in neurofibromatosis type 1 (NF1). Upwards of 80% of individuals with NF1 having some form of neurocognitive dysfunction, resulting in significant morbidities across the lifespan.^1–3^ Research has documented a NF1 cognitive phenotype including downward-shifted intellect, high prevalence of Attention-Deficit/Hyperactivity Disorder (ADHD), executive dysfunction, and visuospatial deficits, some of which mimic learning defects in the NF knockout mouse.^4–7^

Suggested mechanisms underlying cognitive deficits in NF1 include the lack of neurofibromin resulting in overactivity of Ras and hyperactivation of downstream signaling cascades including the Ras/ERK pathway, which is vital in long-term potentiation (LTP) and cortical plasticity involving the hippocampus and medial prefrontal cortex.^8–12^ Trials manipulating the Ras/ERK pathway have demonstrated amelioration of cognitive impairments in NF1 mouse models.^13,14^ Human trials targeting the disrupted pathway appear promising.^1,15^ The deregulation of the Ras/ERK cascade also results in enhanced GABA release, negatively impacting LTP through enhanced inhibition in murine models.^16,17^ Increased GABA release also inhibits prefrontal cortical circuits necessary for working memory, and has been demonstrated in murine *and* human models.^14,18,19^

The most severe neurocognitive dysfunction in individuals with RAS/MAPK pathway disorders, including NF1, appears to be associated with mutations affecting downstream transducers of Ras such as *MEK1* and *BRAF*.^16,20^ This, paired with recent successful clinical trials of *MEK* and *BRAF* inhibitors in attenuating plexiform neurofibromas (PN) and low grade gliomas, has generated interest in the effect MEKi may have on cognition in NF1.^21–25^ Two preclinical trials suggest an impact on cognitive functions with inhibition of MEK in Nf1 mouse models, with conflicting results.^26,27^

The primary aims of this study were to examine changes in memory and executive functions in patients with NF1 on MEKi treatment for PN over the first year of therapy.

## Methods

This was a single-arm, multi-center ancillary cognitive study evaluating changes in cognitive functions in the first year of treatment on a MEKi. Eligible participants were 1. diagnosed with NF1 per NIH criteria or through germline NF1 mutation in a CLIA certified laboratory, 2. enrolled on a clinical trial of a MEKi for the treatment of a PN (including NCT03962543, NCT02096471, NCT02407405, NCT02124772), 3. between the ages of 4 and 35, 4. without significant sensory or motor impairment, and 5. primarily English or Spanish dominant.^28^ This study was approved by the IRB of the PI’s institution and at each of the participating institutions and all participants were consented.

Participants underwent a pre-treatment neurocognitive evaluation. Three additional evaluations were completed over the first year on therapy: at 12 weeks (+/-4 weeks), 24 weeks (+/- 4 weeks), and 48 weeks (+/- 8 weeks). The 48-week assessment was added to the protocol after study initiation resulting in some patients not having 48-week data. Participants taken off treatment early (e.g., for toxicity or lack of response) completed the cognitive assessment at the final study visit, while still taking study drug. At each time point, participants completed the cognitive assessment (Cogstate) and a consistent parent completed a questionnaire of executive functioning (Behavior Rating Inventory of Executive Function; BRIEF). Participants ≥18 completed the self-report BRIEF if an adult caregiver was not present.

All data was managed by the coordinating center (CNH). Individual participants’ data was monitored in real time by the PI (KSW) and findings were conveyed to sites if scores were ≥1.5 SD below the mean, or if there was significant decline on more than two measures over time, allowing for additional monitoring or evaluation for neurotoxicity as needed.

### Study Measures

We were able to study a large, diverse sample with minimal institutional/participant burden using a focused computerized assessment battery (Cogstate) targeting neurocognitive processes most sensitive to change and predicted to be affected by MEKi (i.e., learning/memory, working memory, attention, and processing speed). We also used a complementary rating scale (BRIEF) to evaluate real-world executive functioning. The entire assessment was completed in ≤30 minutes.

#### Cogstate

Cogstate is a computerized testing software package that offers a range of semi-automated assessment modules for individuals aged 4-90 years.^29^ All tasks were developed as computerized adaptations of traditional neuropsychological measures. For this study, we use four tasks in children age six and above (Detection, Identification, One-Card Learning, and One-Back) and two tasks (Detection and Identification) in children ages four and five. These tasks assess specific cognitive functions including visual learning/memory, working memory, attention, and reaction time. The primary performance outcome variables were visual learning/memory (One-Card Learning) and working memory (One-Back).

Cogstate was developed specifically for clinical trials as a sensitive assessment tool that can be administered more frequently than traditional neuropsychological measures and allows for documentation of changes in performance over time, because of a brief practice session, alternative forms, and the ability to collect more data over multiple tests, reducing susceptibility to skewed distributions and floor/ceiling effects.^30,31^ Reliability (intra-class correlation) is 0.77 with good stability and negligible practice effects when testing intervals are greater than one week^32^. For healthy individuals, stability of performance is robust - with test-retest scores differing by approximately 2% on average (traditional measures differ 7-19%)^33^.

#### Behavior Rating Inventory of Executive Function (BRIEF)

The BRIEF^34^ is a widely used behavior rating assessment of executive functioning in everyday life for children aged 3-17 years and adults aged 18-90. The parent-report version consists of 86 items, which map onto 2 broad areas: Behavioral Regulation and Metacognition, as well as two validity scales. The Behavioral Regulation domain is further divided into three clinical subscales: Inhibit, Shift, and Emotional Control, whereas the Metacognition domain is divided into five clinical subscales: Initiate, Working Memory, Plan/Organize, Organization of Materials, and Monitor. The adult self-report is composed of 75 items, which also map onto the two indices of Behavioral Regulation and Metacognition and three validity scales. The two indices are further divided into the same clinical subscales as the parent-report for children, with the exception of the monitor domain being further subdivided into Self-Monitor and Task-Monitor. Parents of children and adult participants are asked to consider the frequency with which each item has been a problem over the last six months, responding on a 3-point Likert scale consisting of “never,” “sometimes,” and “often.” For this study, respondents were instructed to report on observations in the period of time from the previous evaluation rather than over the past six months (except for the pre-treatment assessment), as some of the follow-up assessments on the treatment trials occurred within three months of the previous assessment. Items were developed to be ecologically valid behavioral correlates to presumed neurocognitive difficulties with executive functioning; thus, this measure was selected to provide parent and patient-reported outcomes of problems related to attention, memory, and executive function that occur in everyday life.

Psychometric properties of this measure are strong using normative samples weighted to match ethnic and gender proportions in the US population. Internal consistency is high (alpha range = 0.73 to 0.90) and test-retest reliability exceeds 0.80 for both measures over intervals from 2 to 4 weeks. ^34-36^ Scores are linear transformations of raw scores into T scores (mean = 50, SD = 10) for each of the subscales, indices, and total score; higher scores indicate greater difficulties. As with Cogstate, normative data allows for the computation of age-adjusted scores at any single point in time, and test-retest data provides the information necessary to quantify whether changes in ratings have occurred (declines, no change, or improvements).^37^ The primary BRIEF outcome variables were Behavioral Regulation and Metacognition.

### Statistical Design

Statistical analysis was calculated using IBM SPSS Statistics (Version 26). Changes in performance and ratings over time were evaluated by group-level analysis and individually based reliable change analyses. With both, we quantified whether outcomes changed over time in the sample. We also investigated whether changes in performance differed by age (dichotomized at median of 12 years) and baseline performance (dichotomized as typical or atypical, using 1.5 SD below average).

First, we analyzed group-level change with two sets of dependent samples t-tests, assessing change from pre-treatment to 24 weeks and pre-treatment to 48 weeks. We analyzed the two follow-up time points separately to maximize the available sample at each time point. We calculated effect sizes between mean scores at each pair of visits to estimate the mean amount of change that occurred between time points. To adjust for multiple comparisons, we adjusted critical values within each set of analyses (e.g., 4 t-tests for BRIEF ratings at *p* = .05/4 = .0125).

Second, we analyzed change in outcomes over time on an individual participant level, using Reliable Change Index (RCI) methodology, which generates a confidence interval (CI) that identifies the expected range of change scores in the normative population, using a test’s standard deviation and test-retest reliability. Once the CI cutoff is established, comparisons between the cutoff and an individual’s change score (T_2_ – T_1_) can be made to determine if clinically significant change has occurred. Thus, RCI analysis allows investigation of whether clinically significant, rather than simply statistically significant, change has occurred, and in which individuals. Cogstate provides the within-subject standard deviation (WSD) from the test’s normative sample to calculate RCI, and we calculated metrics for the BRIEF using the test’s normative data in the modified practice-adjusted RCI formula outlined in Chelune.^37, 38^

For each of the outcomes, two-tailed 90% CIs were constructed. In a normative sample, a 90% CI identifies 5% as declined, 90% as stable, and 5% as improved. We chose two-tailed because at this stage of research with MEKi, it is equally important to identify either detrimental (possibly neurotoxic) or positive effects on functioning. Individual change scores were calculated between the pre-treatment and the 24-week, and 48-week evaluations, and classified as “declined,” “stable,” or “improved” on each outcome for each follow-up time point using RCI methodology.

To compare the frequencies of classification between the normative/expected (5/90/5%) and NF groups, we used Chi-square.^39^ Due to the small number of individuals whose scores declined and in order to meet test assumptions regarding expected cell sizes, we collapsed our analyses to a 2 (group: normative/expected, NF) x 2 (change status: declined or stable, improved) Chi-square. This statistical approach answers the question: Does memory or executive functioning improve in a greater proportion of individuals who are taking a MEKi, compared to the proportion expected in the general population who are tested twice? We hypothesized that a greater proportion of individuals receiving a MEKi would change in performance compared to normative expectations. We adjusted these critical values for multiple comparisons as well (*p*_critical_ = .0125).

Third, to assess the influence of age and baseline performance level on RCI-based classification, we used a set of 2 (age: ≤12 years, >12 years; baseline performance level: typical, atypical) x 2 (change status: declined/stable, improved) Chi-square tests. This allowed us to test whether older or younger children were more likely to change, and whether those with atypical scores at baseline were more likely to change than those with typical baseline performance.

Given the exploratory nature of these additional analyses, we did not adjust p-values for multiple comparisons.

## Results

Seventy-four patients ages five to 27 years enrolled on the study. One patient was ineligible for the treatment study, one declined participation, one did not complete the measures at pre-treatment, and five discontinued treatment prior to the 24-week time point. Seven discontinued treatment by the 48-week time point, and twelve patients were outside the 48-week test window when this assessment was amended to the protocol. In total, 59 patients completed the pre-treatment and 24-week assessments, 40 (68%) of whom also had 48-week data. The median age of participants was 12 years (5-27), 64% were male, and 68% were Caucasian. Most participants on this study were treated with Selumetinib, with the remainder treated with Mirdametinib or Trametinib (Table 1).

**Table 1.** Patient and Treatment Characteristics.

|  |  |  |
| --- | --- | --- |
| Age in years | Mean (SD) | 12.66 (5.74) |
|  | Median | 12 |
|  | Range | 5-27 |
| Gender (Males) | N (%) | 38 (64%) |
| Race/Ethnicity |  | <u>N (%)</u> |
|  | Caucasian | 40 (68%) |
|  | African American | 6 (10%) |
|  | Asian | 3 (5%) |
|  | Multiple | 5 (8%) |
|  | Hispanic | 3 (5%) |
|  | Other | 2 (3%) |
| Drug |  | <u>N (%)</u> |
| *Selumetinib (NCT02407405)* |  | 54 (92%) |
| *Mirdametinib (NCT03962543, NCT02096471)* |  | 4 (7%) |
| *Trametinib (NCT02124772)* |  | 1 (1%) |

### Patient/Observer Reported Outcomes (BRIEF)

Descriptive statistics of the 52 BRIEF parent and seven adult self-reports are provided in Table 2. Dependent samples t-tests indicated small but significant improvements in BRIEF scores from pre-treatment to 24-week follow-up (MCI: *t*_(58)_ = 2.41, *p* = .02, Cohen’s *d* = 0.18; BRI: *t*_(58)_ = 3.03, *p* = .004, *d* = 0.24) and 48-week follow-up (MCI: *t*_(39)_ = 2.70, *p* = .01, *d* = 0.27; BRI: *t*_(39)_ = 2.37, *p* = .02, *d* = 0.26); only the MCI analyses were significant after correcting for multiple comparisons (*p* < .0125). RCI analyses indicated that very few individuals treated with a MEKi declined, with distribution at the 24-week assessment on MCI being 5/80/15% and on BRI 2/85/13% for declined/stable/improved, respectively. After collapsing the declined and stable categories, the 24-week distributions were not found to be different from the expected collapsed distribution of RCI classifications in the normative population (95/5%); MCI: 85/15%, *Chi Square* = 4.86, *p* = .03, OR = 3.4; BRI: 87/13%; *Chi Square* = 3.49, *p* = .08, OR = 2.9. The proportions were significantly different at 48 weeks for MCI (75/25%; *Chi Square* = 11.95, *p* = .001, OR = 6.3; see Figure 1), such that the proportion classified as “improved” was larger than expected, but not for BRI (85/15%; *Chi Square* = 3.95, *p* = .08, OR = 3.4).

**Table 2.** Mean Cognitive Scores from Pre-Treatment to 48-week Follow-Up.

|  | Pre-Treatment |  |  | 24-week |  | 48-week |  |
| --- | --- | --- | --- | --- | --- | --- | --- |
|  | M (SD) | Median (Min-Max) | N (%) Impaired | M (SD) | Median (Min-Max) | M (SD) | Median (Min-Max) |
| BRIEF MCI (Tscore) | 54.93 (10.95) | 55.0 (36-84) | 11 (19%) | 52.95 (11.07) | 53.0 (34-82) | 51.7 (10.6) | 51.5 (31-71) |
| BRIEF BRI (Tscore) | 51.59 (10.31) | 51.0 (35-79) | 6 (10%) | 49.1 (9.66) | 48.0 (35-81) | 48.73 (9.36) | 46.5 (35-68) |
| Cogstate ONB Speed (z-score) | -0.24 (1.42) | -0.1 (-3.7-2.82) | 9 (15%) | -0.18 (1.33) | 0.1 (-4.4-2.89) | -0.35 (1.11) | -0.3 (-3.11-2.89) |
| Cogstate OCL Accuracy (z-score) | -0.22 (1.37) | -0.4 (-2.67-3.0) | 8 (14%) | -0.14 (1.57) | 0.0 (-5.78-3.78) | -0.04 (1.49) | -0.2 (-3.3-3.0) |

*M = Mean, SD = standard deviation*
*Note: BRIEF N=59 at pre-treatment and 24 weeks, N=40 at 48 weeks; Cogstate N=57 at pre-treatment and 24 weeks, N=39 at 48 weeks); some individual sample sizes vary*
*\*Impaired scores were defined as $\geq 1.5$ SDs below the mean*

**Figure 1.**
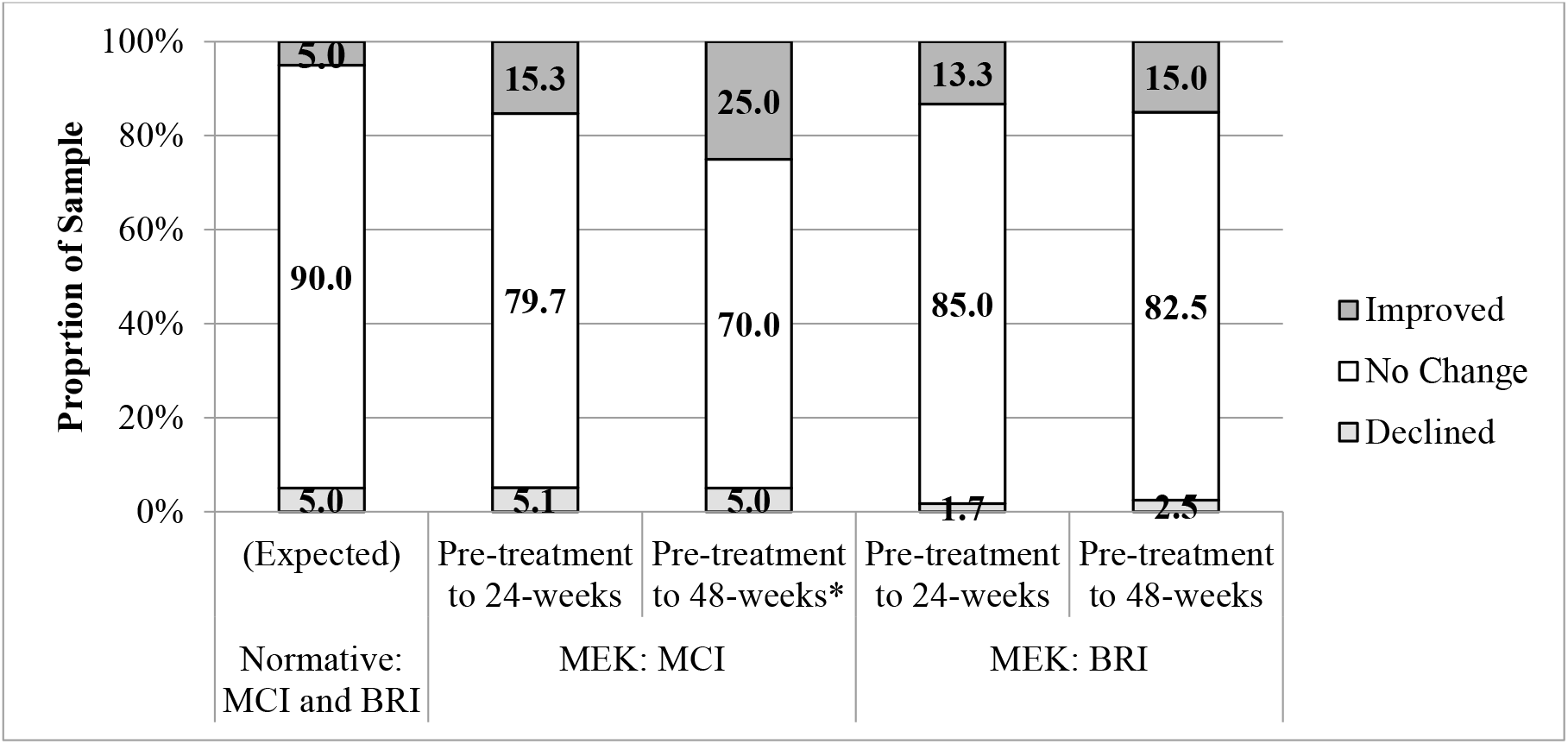
RCI-based outcomes on observer-rated executive functions following 24- and 48-weeks of treatment. BRIEF MCI = Behavior Rating Inventory of Executive Functioning, Metacognition Index BRIEF BRI = Behavior Rating Inventory of Executive Functioning, Behavioral Regulation Index RCI = Reliable Change Index MEK = Received Mitogen-activated protein kinase inhibitor *Significantly different from Normative (Expected) proportions using 2-tailed 90% CI

When examining RCI-based changes in executive function ratings over time in the subsample that completed all assessments (n=40), overall declines in executive function ratings were seen in only 5% of the sample on the MCI and 2.5% on the BRI by 48-weeks. Most of the sample showed no net changes: 70% on MCI and 88% on BRI (either declined/improved at 24-weeks but then returned to baseline, or never changed). Of the 25% who showed 48-week improvement on MCI, half had improved by 24-weeks and then were stable, and the other half were stable at 24-weeks but improved at 48-weeks. Of the 15% who showed overall improvement on BRI, 5/6 improved at 24-weeks and were then stable, while one participant was stable at 24-weeks then improved at the final follow-up. Many of the participants who were stable at 24-weeks had enrolled in the study prior to the addition of the 48-week assessment (MCI n=17; BRI n = 15), as were some who had improved at 24-weeks (MCI n=2; BRI n=5) limiting the ability to assess long-term improvement in these individuals.

### Primary Performance Outcomes (Cogstate)

Analysis of change in Cogstate working memory (ONB Speed) and visual learning/memory (One-Card Learning) did not reveal significant change in performance from pre-treatment to 24- or 48-week follow-up (ONB Speed: *t*_(55)_ = -0.71, *p* = .48, Cohen’s *d* = 0.08; *t*_(40)_ = 0.70, *p* = .49, *d* = -0.08, respectively; One-Card Learning: *t*_(54)_ = -0.48, *p* = .64, *d* = 0.08; *t*_(1,40)_ = 0.05, *p* = .96, *d* = -0.01, respectively). Again, RCI analyses indicated that few individuals treated with a MEKi declined, particularly on the OCL task, with distribution at the 24-week assessment on ONB being 14/66/20% at 24 weeks and 12/66/22% at 48 weeks, and on OCL 5/87/7% at 24 weeks and 0/98/2% for declined/stable/improved, respectively. After collapsing the declined and stable categories, the ONB Speed distributions were different from the expected collapsed distribution of RCI classifications in the normative population (95/5%) at the 24-week assessment (80/20%; *Chi Square* = 8.36, *p* = .004, OR = 4.6) and the 48-week assessment (78/22%; *Chi Square* = 9.34, *p* = .004, OR = 5.3). The distributions of RCI classifications for OCL accuracy were not significantly different at either the 24-week assessment (93/7%; *Chi Square* = 0.34, *p* = .72, OR = 1.5) nor at the 48-week assessment (98/2%; *Chi Square* = 0.47, *p* = .67, OR = 0.5); see Figure 2.

**Figure 2.**
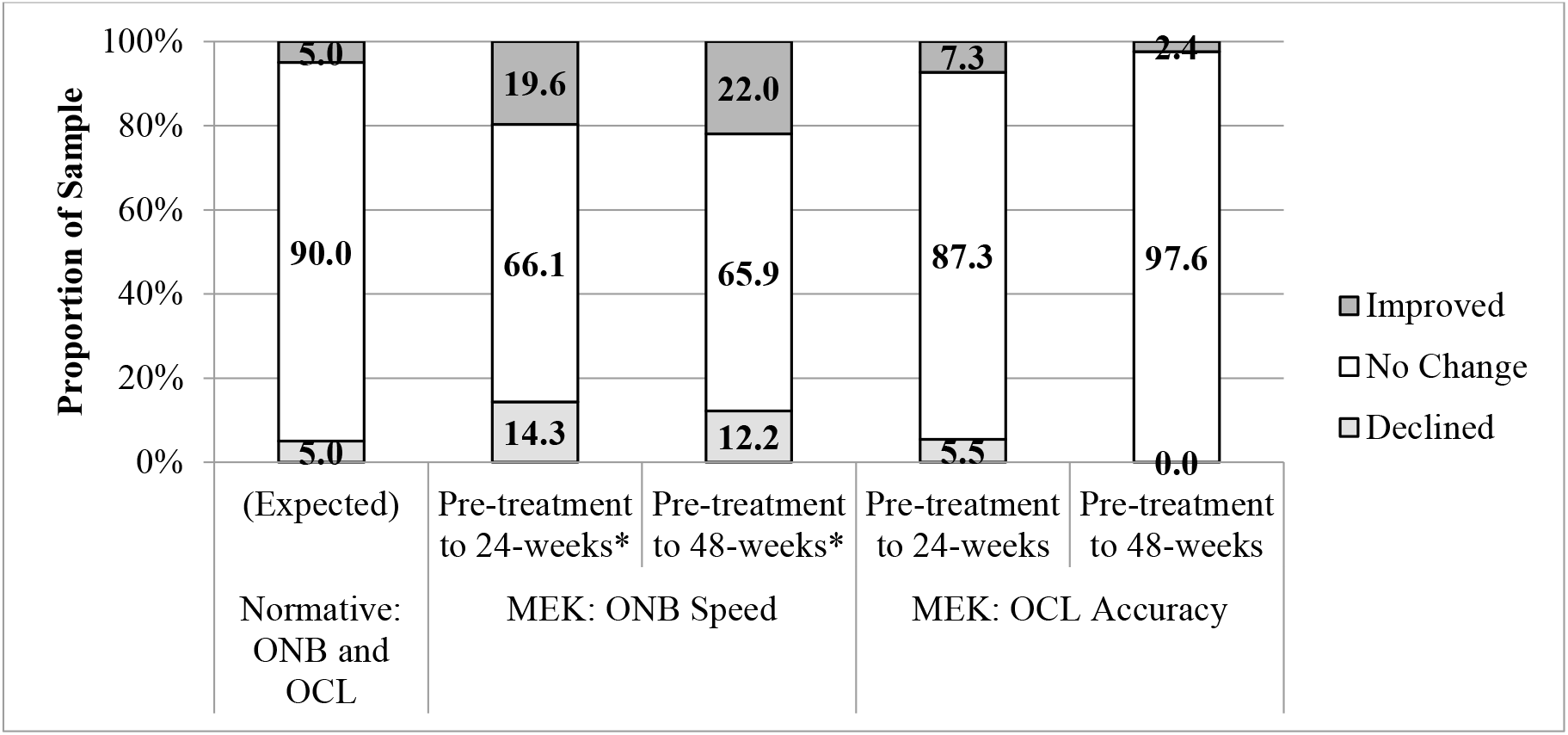
RCI-based outcomes on performance-based learning and working memory following 24- and 48-weeks of treatment. ONB = One-Back Task OCL = One-Card Learning Task RCI = Reliable Change Index MEK = Received Mitogen-activated protein kinase inhibitor *Significantly different from Normative (Expected) proportions using 2-tailed 90% CI

**Figure 3.**
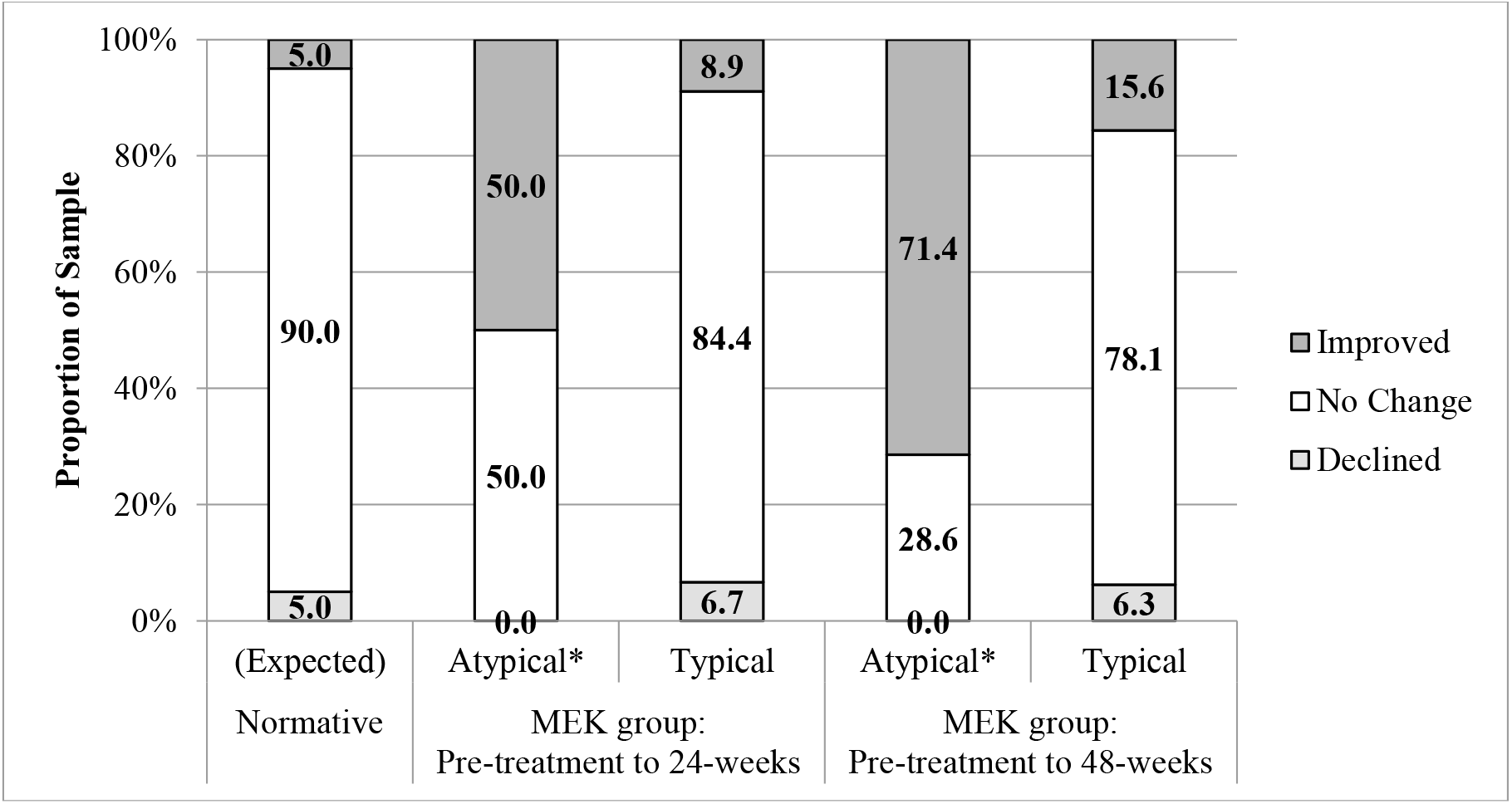
RCI-based change in observer-rated executive function outcomes on the BRIEF MCI associated with baseline performance levels. BRIEF MCI = Behavior Rating Inventory of Executive Functioning, Metacognition Index RCI = Reliable Change Index MEK = Received Mitogen-activated protein kinase inhibitor *Significantly different from the corresponding Not Impaired group and Normative (Expected) proportions using 2-tailed 90% CI

**Figure 4.**
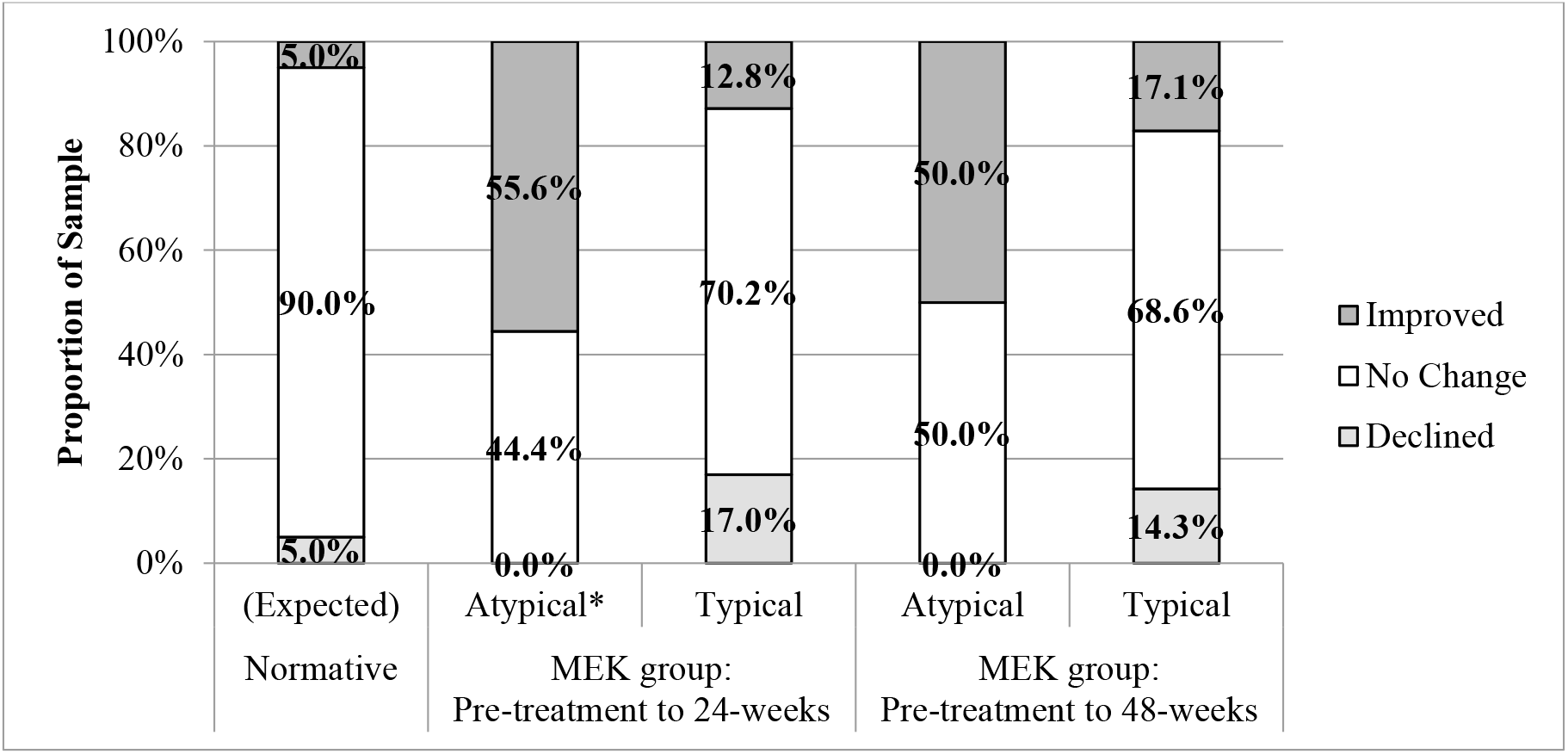
Performance-based working memory outcomes (ONB Speed) associated with baseline performance levels. ONB = One-Back Task RCI = Reliable Change Index MEK = Received Mitogen-activated protein kinase inhibitor *Significantly different from the corresponding Not Impaired group and Normative (Expected) proportions using 2-tailed 90% CI

When examining RCI-based changes in performance over time in the subsample that completed all assessments, overall declines were rare (12% of the sample on ONB and 0% on OCL). Most of the sample showed no net changes: 66% on One-Back and 97% on One-Card Learning. Of the nine participants who showed improvement on ONB by 48-weeks, 1/3 (n=3) had improved by 24 weeks and then were stable, the remaining 2/3 (n=6) did not improve until 48-weeks. As with the executive functioning ratings, many of those who had stable or improved performance at 24-weeks did not have 48-week data (n = 8 who were stable, 5 who had improved on ONB; n = 13 stable and 1 improved on OCL), limiting the possibility of assessing long-term gains in these participants.

### Predictors of RCI Classifications

#### Age

To determine if the age of the participant related to outcome, we compared children ≤12 years (younger cohort, n=34) to adolescents and young adults ≥13 (n=25). There were no differences in the proportion of participants whose scores declined or did not change (collapsed categories) vs improved on BRIEF or Cogstate at either 24- or 48-week follow-ups (all *p* > .10).

#### Baseline level of functioning

The contribution of pre-treatment functioning to outcomes was examined. In the normative (test-retest) BRIEF sample, there were no associations of initial rating level (atypical vs typical) and change in score at retest (p > .05). Within the MEKi sample, we found that participants with baseline MCI ratings in the impaired (atypical) range (≤ 1.5 standard deviations below the mean, which was seen in n=11; 19%) were more likely to show significant improvement over the course of treatment. At 24 weeks, 46% of the impaired pre-treatment group showed significant improvement, compared to only 8% of the not-impaired group (*Chi Square* = 9.54, *p* = .008, OR = 9.22). Similarly, at the 48-week follow-up, participants with impaired scores at pre-treatment were significantly more likely to show meaningful improvement on MCI (63%) than those whose initial scores were not impaired (16%; *Chi Square* = 7.50, *p* = .02, OR = 9.0).

A similar pattern was seen on BRI between those impaired (*n =* 6) versus not impaired at pre-treatment; however, the proportions were not as striking, and the comparisons were not statistically significant. A 24-weeks, 33% of those with impaired pre-treatment scores improved compared to 9% of those with non-impaired pre-treatment ratings (*Chi Square* = 2.94, *p* = .14, OR = 4.8). At 48-weeks, 25% of those with impaired pre-treatment scores improved (again, not significantly so) compared to 14% of those with non-impaired pre-treatment ratings (*Chi Square* = 0.35, *p* = .49, OR = 2.1).

Using the same cutoff criteria of 1.5 SD below the mean, statistical significance was reached on one of the Cogstate tasks from pre-treatment to follow-up. At 24-weeks, significant improvement on ONB Speed was demonstrated in 56% of those with impaired performance at pre-treatment (*n* = 9), and only 13% of those with non-impaired pre-treatment performance (*Chi Square* = 8.76, *p* = .01, OR = 8.5). At 48-weeks, differences were not significant, though the pattern was similar: 50% of those performing in the impaired range at pre-treatment improved while only 17% of the participants with non-impaired pre-treatment performance showed improvement (*Chi Square* = 3.78, *p* = .09, OR = 4.8).

Neither analysis on Cogstate One-Card Learning (accuracy) was statistically significant. Eight patients had impaired OCL performance at pre-treatment. At both 24- and 48-weeks, the only participants who improved were those with non-impaired pre-treatment performance, and chi-square tests were not significant (24-week *Chi Square* = 0.73, *p* = .99, OR 0.0; 48-week *Chi Square* = 0.51, *p* = .99, OR = 0.0).

## Discussion

The recent MEK inhibitor trials have provided a key opportunity to enhance our understanding of possible mechanisms of action in NF1-associated cognitive impairment, as well as to accelerate our progress toward finding effective therapeutics with the potential for improving these neurocognitive deficits in patients. Our innovative methodology of developing an ancillary cognitive study that could be carried out across multiple ongoing MEK inhibitor clinical trials allowed us to assess a key question in NF1 research and maximize enrollment without having to design and recruit to a separate clinical trial. The methodology we applied showed excellent feasibility in a multi-center trial, supporting the use of these or similar methods in future cognitive trials in NF. This study is the first to investigate the effects of MEK inhibitors on cognitive functioning in individuals with NF1 and provides novel information to the field. Given the mixed results of the NF1 murine model research regarding MEK inhibition and cognition/development, the most important finding of this study is the lack of evidence of neurotoxicity within the first 48-weeks of therapy in individuals with NF1 ages five years and older.^26,27^ Performance scores of visual learning remained stable for the majority of participants. Similarly, performance on a working memory task and symptom ratings of both metacognitive and behavioral regulation aspects of executive function showed stability or improvement over the course of their treatment trial. We were unable to evaluate for potential neurotoxicity in children under the age of five, and this will be an important next step in this research given the greater susceptibility associated with rapid brain development and plasticity in this early period.^40^

Group-level analyses of change indicated some small but significant improvements in parent/patient-reported executive functioning but not performance-based tasks of executive function. By using a Reliable Change Index framework, however, our statistical approaches allowed for an in-depth and clinically meaningful assessment of individual outcomes.

Based on parent/patient-reported symptom ratings of everyday executive functions, results suggest clinical improvement of metacognitive regulation functions observable by 48-weeks or sooner in a larger proportion of participants receiving treatment. The age of the participant did not impact these outcomes, at least when comparing results in school-age children to adolescents/young adults. However, pre-treatment rating of executive dysfunction did relate to greater improvements in metacognitive and behavioral regulation functions than in participants rated as unimpaired prior to treatment. This finding was most prominent for metacognitive functions with significant improvements reported by the 24-week follow-up with additional improvements rated at the 48-week assessment. While it is possible that regression to the mean played a role in these findings, the marked pattern we saw in patients treated with a MEKi was not seen in the BRIEF normative sample.

On performance-based measures, a significant proportion of the treatment sample showed clinical improvement on a working memory task over the first 24- to 48-weeks on therapy, and again there was no relationship with age, but those with greater pre-treatment impairment had greater improvements at 24-weeks than in participants with non-impaired performance prior to treatment. In contrast, there were not notable improvements in visual learning/memory.

Results of parent/patient-reported outcome measures were more robust than findings on performance-based measures. It is common for these assessment approaches to produce unique information about patient functions that are complementary.^37, 41^ There exists the possibility that there is some inherent bias in parent and patient reports of functioning with some expectation of improvement. However, the cognitive study was presented with non-directional hypotheses, which should minimize the issue of such bias. Additionally, parents and patients completed new forms at each assessment blind to their previous ratings.

It is also possible that the parent/patient report captured a broader improvement in functioning related to decreased pain and increased mobility or other functions directly related to changes in the PN. The patients on the SPRINT trial (NCT02407405) were enrolled based on the severity of their PN and associated pain, disfigurement, and functional limitations. In addition to tumor response, the patients in that study showed clinically meaningful improvement in pain intensity, pain interference, and overall quality of life.^21^ Therefore, it is possible that these improvements may have resulted in more engagement in school, social, and other settings that were essentially captured by the BRIEF. However, as many of the cases in the SPRINT trial showed improvement in pain and quality of life earlier in the course of treatment (within the first 12 weeks), we may be discovering unique cognitive improvements as well. While parent/patient report of improvement was often first seen at 24-weeks, many of the performance-based cognitive improvements (when present) were not evident until 48-weeks.^21^ This may suggest that there is a true association between treatment with a MEK inhibitor and improved cognitive functions that do not emerge until closer to a year on therapy.

This study was an ancillary to several single-arm, open label clinical trials, which limits our ability to compare and contrast these findings to a comparison group (treated or untreated), which is an obvious limitation to this research. The use of RCI partially accounts for this, as it uses data from normative test-retest samples as comparison. Future plans include analyzing the relationship between cognitive, pain, and mobility outcomes to determine if these factors are related to reported improvement in executive function or if symptom reports of everyday executive functions are more sensitive measures of cognitive change in the context of treatment with a MEK inhibitor. Our data cannot speak to the impact treatment beyond one year may have on cognition or the durability of these findings. However, a randomized trial of Selumetinib versus carboplatin/vincristine in patients with NF1 and low-grade glioma has recently been initiated (NCT03871257), which will address the limitations related to a lack of comparison group, durability, and a general enhancement of our interpretation of these initial findings.

In conclusion, cognitive impairments and learning disabilities in NF1 are a significant life-long morbidity and the lack of effective treatments continues to pose a significant unmet need. This study provided a first investigation into potential mechanisms of action related to cognitive dysfunction in NF1 involving the RAS/MAPK pathway and provides important initial findings regarding the effects of MEK inhibitors on cognitive function. First and foremost, the results of this study do not indicate any significant cognitive deterioration in functioning over the first 48-weeks of treatment with a MEK inhibitor that might suggest drug neurotoxicity. Our data shows evidence of real-world functional and clinical improvement in executive functioning and improvement on a working memory task that emerge by around 24 weeks on a MEK inhibitor and with continued improvement up to 48 weeks of treatment, particularly for patients with baseline cognitive dysfunction, as would be expected. There is enough preliminary support of possible benefit of MEK inhibitors on cognitive functioning in NF1 that future research in this area should be a focus for the NF1 community.

## Data Availability

This data is not currently openly available.

## Acknowledgements

This research was supported by the Children’s Tumor Foundation, the Jennifer and Daniel Gilbert Neurofibromatosis Institute, and, in part, by the Intramural Research Program of the Center for Cancer Research, NCI, NIH, Bethesda, MD. This project has been funded in whole or in part with federal funds from the National Cancer Institute, National Institutes of Health, under Contract NO. 75N91019D00024, Task Order No. 75N91019F00129. The content of this publication does not necessarily reflect the views or policies of the Department of Health and Human Services, nor does mention of trade names, commercial products, or organizations imply endorsement by the U.S. Government. None of the funding sources had any role in the writing of the manuscript or the decision to submit.

## Author Contributions

Karin S. Walsh contributed to the overall study design, literature search, data collection, data analysis, data interpretation, creation of figures, and writing

Pamela L. Wolters contributed to the study design, data collection, data interpretation, and writing

Brigitte C. Widemann contributed to the study design, data collection, data interpretation, and review of the manuscript

Allison del Castillo contributed to the study design, literature search, data collection, data analysis, data interpretation, and writing

Maegan D. Sady contributed to data analysis, data interpretation, creation of figures, and writing Tess Inker contributed to the literature search, data collection, and review of the manuscript Marie Claire Roderick contributed to data collection and review of the manuscript

Staci Martin contributed to data collection, data interpretation, and review of the manuscript Mary Ann Toledo-Tamula contributed to data collection and review of the manuscript

Kari Struemph contributed to data collection and review of the manuscript Iris Paltin contributed to data collection and review of the manuscript

Victoria Collier contributed to data collection and review of the manuscript

Kathy Mullin contributed to data collection and review of the manuscript Michael J. Fisher contributed to data collection and review of the manuscript

Roger Packer contributed to data collection and review of the manuscript

Potential Conflicts of Interests

Karin S. Walsh declares no conflict of interest

Pamela L. Wolters declares no conflicts of interest

Brigitte C. Widemann declares no conflicts of interest

Allison del Castillo declares no conflicts of interest

Maegan D. Sady declares no conflict of interest

Tess Inker declares no conflict of interest

Marie Claire Roderick declares no conflict of interest

Staci Martin declares no conflicts of interest

Mary Ann Toledo-Tamula declares no conflict of interest

Kari Struemph declares no conflict of interest

Iris Paltin declares no conflict of interest

Victoria Collier declares no conflict of interest

Kathy Mullin declares no conflict of interest

Michael J. Fisher declares no conflict of interest

Roger Packer declares no conflict of interest

